# Second-generation optoacoustic imaging of breast cancer patients

**DOI:** 10.1101/2021.10.15.21264936

**Authors:** Jan Kukačka, Stephan Metz, Christoph Dehner, Alexander Muckenhuber, Korbinian Paul-Yuan, Angelos Karlas, Ernst Rummeny, Dominik Jüstel, Vasilis Ntziachristos

**Affiliations:** Helmholtz Zentrum München (GmbH), Neuherberg, Germany, Institute of Biological and Medical Imaging; Helmholtz Zentrum München (GmbH), Neuherberg, Germany, Institute of Computational Biology; Technical University of Munich, Germany, School of Medicine, Chair of Biological Imaging at TranslaTUM; Technical University of Munich, Germany, Department of Diagnostic and Interventional Radiology; Technical University of Munich, Germany, Institute of General and Surgical Pathology; Klinikum rechts der Isar, Munich, Germany, Clinic for Vascular and Endovascular Surgery

**Keywords:** Multispectral optoacoustic tomography, In vivo imaging, Breast cancer, Tumor-associated microvasculature, Image quality enhancement, Ultrasound

## Abstract

Since the initial breast transillumination almost a century ago, breast cancer imaging using light has been considered in different implementations aiming to improve diagnostics, minimize the number of available biopsies, or monitor treatment. However, due to strong photon scattering, conventional optical imaging yields low resolution images, challenging quantification and interpretation. Optoacoustic imaging addresses the scattering limitation and yields high-resolution visualization of optical contrast, offering great potential value for breast cancer imaging. Nevertheless, the image quality of experimental systems remains limited due to a number of factors, including signal attenuation with depth and partial view angle and motion effects, particularly in multi-wavelength measurements. We developed data analytics methods to improve the accuracy of handheld optoacoustic breast cancer imaging, yielding second-generation optoacoustic imaging performance operating in tandem with ultrasonography. We produced the best images yet with handheld optoacoustic examinations of the human breast and breast cancer, in terms of resolution and contrast. Using these advances, we examined optoacoustic markers of malignancy, including vasculature abnormalities, hypoxia, and inflammation, on images obtained from breast cancer patients. We achieved the best optoacoustic images of the human breast ever obtained using handheld examination, advancing the diagnostic and theranostic potential of the hybrid optoacoustic-ultrasound (OPUS) examination over routine ultrasonography.

## 1 Introduction

Imaging of optical contrast has been utilized in breast cancer applications since 1931,^1^ but is inherently challenging due to light absorption and scattering in tissue. Resolving optical contrast enables imaging of endogenous tissue chromophores or external contrast agents, allowing visualization of pathophysiological hallmarks of breast cancer *in vivo*. These include structural changes (e.g. angiogenesis^2^), physiological parameters (e.g. hemoglobin concentration and oxygen saturation^3-5^), and gene expression (using probes targeting overexpressed receptors^6^ or probes activated by tumor-associated enzymes^7^). The ability to assess these additional tumor features could reduce unnecessary biopsies,^8-10^ aid in the diagnosis of breast cancer, lead to more efficient treatment planning and personalized medicine,^11,12^ and support the monitoring of therapies^13-15^ and drug development.^12^ Moreover, characterization of oxy- and deoxy-hemoglobin in the near-infrared range has been shown to have predictive value for a patient’s response to chemotherapy.^13,16-21^

Key limitations of conventional optical imaging include low spatial resolution and quantification challenges due to the strong photon scattering of light in tissue. These challenges reduce the accuracy and sensitivity of optical examinations. The use of externally administered agents, including indocyanine green,^22^ nanoparticles,^23^ or fluorescent agents that target enzymes and over-expressed receptors^7^ has been considered to improve the performance of examinations. Nevertheless, the administration of contrast agents can be problematic in diagnostic protocols due to safety regulations and pharmacokinetics or photo-stability concerns.^24,25^ The low resolution and limitations in regard to the use of contrast agents continue to limit the broad use of optical imaging approaches in mainstream breast radiology.

Optoacoustic (OA) imaging addresses the photon scattering limitations of conventional optical imaging methods. In OA imaging, ultrasound (US) waves are detected, which are produced in response to transient illumination absorbed by tissue chromophores. The US waves captured on the tissue surface are mathematically combined in a tomographic scheme that can produce images of optical absorption in tissue with a resolution dictated by ultrasound diffraction. By detecting optical absorption using US waves, OA imaging is insensitive to photon scatter and yields images with higher resolution and fidelity than diffuse optical imaging.^26,27^ For these reasons, OA imaging is better suited for breast cancer examinations than conventional optical imaging approaches. The method has already been considered since the mid-90s^28,29^ and has demonstrated the feasibility of imaging endogenous optical contrast deep in breast tissue *ex vivo*^2^ and *in vivo* using a handheld system.^30^ However, these initial OA prototypes employed detectors with an overall low number of ultrasound elements (12 and 32, resp.) and simple inversion techniques, typically based on back-projection algorithms. Since then, several different implementations have been considered for *in vivo* breast cancer imaging, with attention directed toward bed-based systems using planar detector arrays,^31-33^ circular detector arrays,^34,35^ and hemispherical detector arrangements.^36,37^ Bed-based systems have stationary geometries and immobilize the scanned breast to avoid motion artefacts. Immobilization of the breast also enables 3D scanning and the use of large apertures, leading to image quality improvements and minimized intra-operator variability.^38^ Nevertheless, bed-based scanners are limited by the cost and difficulty of interfacing a complex 3D scanning geometry to the human breast. Moreover, their integration into the existing breast examination workflow may be challenging since dedicated scanners require separate examinations. Therefore, despite the attention given to bed-based scanners, handheld systems can offer ubiquitous OA examination, especially since they can be seamlessly integrated with handheld ultrasonography, which is routinely employed in clinical breast examinations, adjunct to x-ray mammography.^39-41^ US and OA utilize different contrast mechanisms, and thus capture complementary morphologic and functional features of a tumor and the surrounding tissue that could enhance the performance of the examination, while fitting seamlessly into today’s clinical workflow.

Handheld optoacoustic and ultrasound (OPUS) imaging has been demonstrated by two prominent systems. The *Imagio®* scanner (Seno Medical Instruments Inc.; Texas, USA) uses a linear transducer array with 128 piezo elements and a pair of lasers delivering light at two wavelengths (755 nm and 1064 nm), providing co-registered OA and US images.^42^ This scanner was employed in two large, multi-center studies, for adjustment of grading of suspicious breast lesions via predefined semi-quantitative OA features^9,43^ and identification of cancer subtypes.^44^ The *Acuity Echo*® multi-spectral optoacoustic-ultrasound (2G-OPUS) scanner (iThera Medical GmbH, Germany), employed herein, features a curvilinear transducer array with 256 piezo elements and a 145° angular coverage. The curvilinear design affords superior imaging quality compared to linear arrays.^45^ Moreover, using a fast-tunable laser, the *Acuity* scanner enables the collection of 28 images at different wavelengths within 1.1 seconds, offering high spectral definition in the 680–980 nm wavelength range while minimizing motion artifacts. Variants of the *Acuity* scanner have been successfully utilized in clinical studies of melanoma metastatic status,^46^ Crohn’s disease,^47^ brown fat metabolism,^48,49^ normal vasculature^50^ and vascular malformations,^51^ thyroid disease,^52^ systemic sclerosis,^53^ and Duchenne muscular dystrophy.^54^ In two pilot breast cancer studies, multi-spectral optoacoustic tomography (MSOT) revealed increased vascularization in the periphery of tumors and a concomitant reduction in the tumor core, as well as heterogeneous total blood volume and irregular deoxy-hemoglobin (Hb) and oxy-hemoglobin (HbO_2_) signal patterns in the tumor area.^4,55^

Despite the state-of-the-art hardware specifications of the *Acuity Echo®* scanner, which features a curvilinear detector and fast wavelength switching ability, the image quality of handheld scanners is challenged by the limited view angle, which reduces the resolution and contrast achieved and introduces imaging artifacts.^56,57^ In the current work, we aimed to reach the next level of image quality and accuracy in handheld OA imaging by advancing image formation methods. In particular, we hypothesized that using a precise forward model using speed of sound correction, total impulse response correction, and compounding of motion-corrected frames could lead to the most accurate image quality ever achieved in handheld OA imaging of breast cancer. Using this second-generation OPUS (2G-OPUS), we were in particular interested to understand the size and contrast of the smallest structures that can be reliably resolved. For this reason, we further explored dual-band visualization, which separates signals from two frequency bands and improves visualization of fine details over strong background signals. We analyzed data from 22 patients and present a detailed analysis of eight representative cases to better outline the breast cancer features resolved with 2G-OPUS. We showcase both anatomical and spectral features from selected regions of the image. The techniques and findings presented herein highlight, to the best of our knowledge, the most advanced state of handheld breast cancer OA imaging performance.

## 2 Results

### 2.1 Going beyond the state-of-the-art on image quality

Figure 1 shows results from a patient in their 70*’*s with an invasive lobular carcinoma (dashed line). Ultrasonography (Fig. 1A) revealed a large tumor mass approximately 1.5 cm under the surface (arrow), which is marked by a dashed line in all subsequent optoacoustic images. The US image is greatly enhanced by vascular structures (Fig. 1B) upon superimposing the OA image (in red) on the US image (grayscale). The superimposed OA image is the output of the image formation pipeline developed herein and described below.

**Figure 1.**
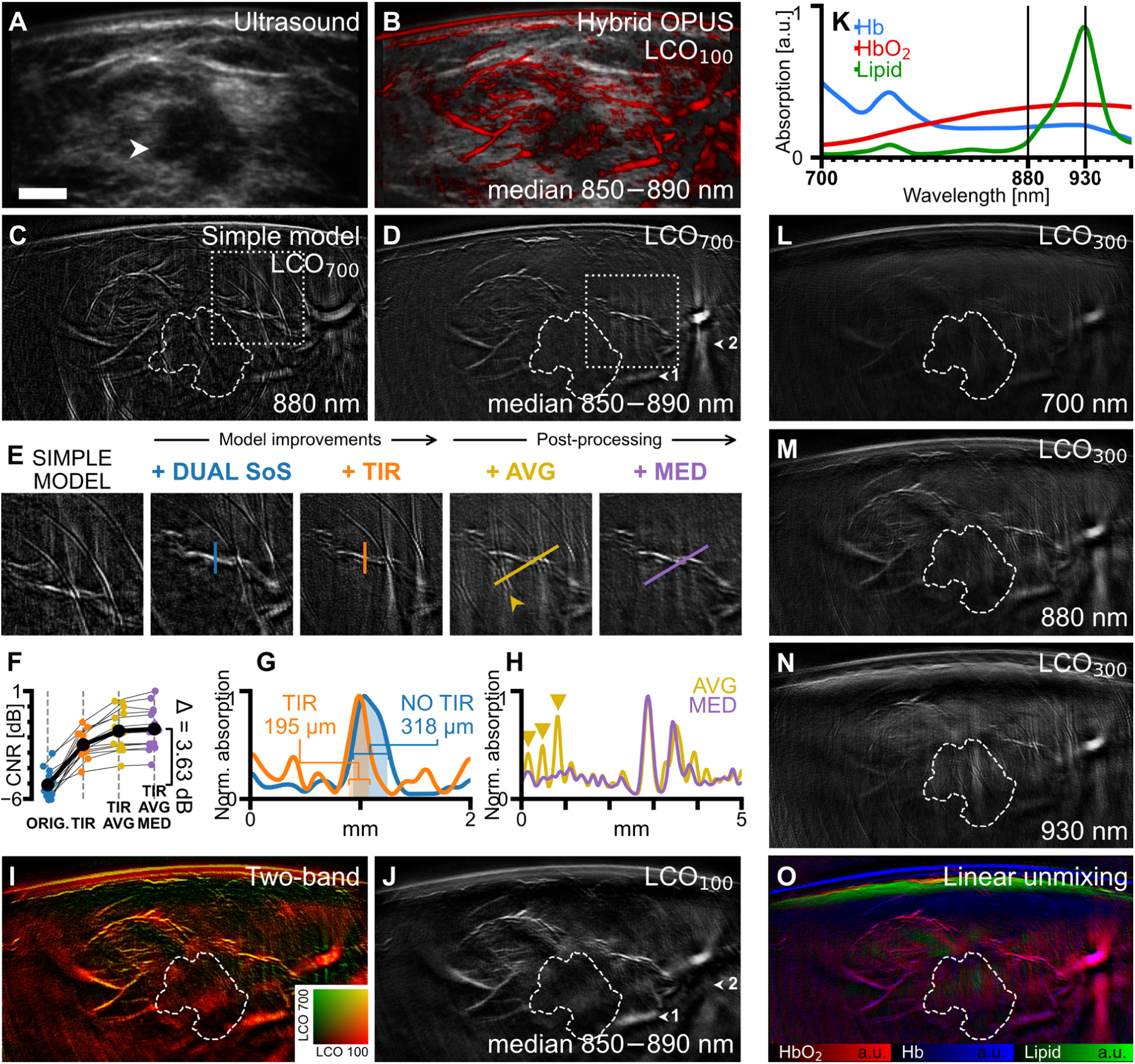
Improvements in image quality and visualization of information contained in 2G-OPUS images demonstrated on a case of invasive lobular carcinoma (Case 1). **A**, Grayscale ultrasound image with hypoechoic tumor mass (white arrow). The tumor core is delineated by dashed contours in subsequent panels. **B**, Hybrid OPUS visualization providing complementary morphologic and functional information about the lesions by overlaying optoacoustic signal (red, LCO_100_, median 850–890 nm) over grayscale ultrasound. **C**, Optoacoustic (OA) image (λ = 880 nm) reconstructed using a simple, uniform speed-of-sound (SoS) model without total impulse response (TIR) correction. **D**, OA image (median 850–890 nm) using band-pass filter with a 700 kHz lower cutoff frequency (LCO_700_). Arrows highlight differences between LCO_700_ and LCO_100_ (shown in J); arrow 1 marks an out-of-plane large blood vessel that is suppressed by LCO_700_, while arrow 2 marks streak artifacts caused by limited view that are prominent in the LCO_700_ variant. **E**, Cut-out marked in C, D showing the cumulative effect of using dual SoS, TIR correction, frame averaging (AVG), and spectral median correction (MED). **F**, Contrast-to-noise ratio (CNR) evaluated in all scans from our study. Mean CNR of images reconstructed without proposed improvements is –5.09 dB, with TIR –2.47 dB, with TIR and AVG –1.59 dB, and with TIR, AVG, and MED –1.46 dB. **G**, Line profiles (normalized to maximum) of a blood vessel cross-section, marked in the panel E, images 2 and 3. Decrease of full width at half maximum from 318 μm (No TIR) to 195 μm (TIR) indicates improved resolution. **H**, Line profiles (normalized to maximum) along the lines marked in the panel E, images 4 and 5. Peaks marked by yellow arrows correspond to ring-shaped artifacts caused by electrical noise (also marked by yellow arrow in E). Suppression of these peaks on the purple curve shows that the spectral median filter removes this type of noise. **I**, Dual-band visualization, obtained by combining image variants using band-pass filters LCO_100_ (J) and LCO_700_ (D) into a single image. **J**, OA image (median 850–890 nm) using band-pass filter with 100 kHz lower cutoff frequency (LCO_100_). **K**, Absorption spectra of deoxy-hemoglobin (Hb), oxy-hemoglobin (HbO_2_), and lipids in the measured range.^60^ **L, M, N**, OA images (LCO_300_) at 700 nm, 880 nm, and 930 nm, respectively, representing the strongest relative absorption of Hb, HbO_2_, and lipids. **O**, Concentrations of HbO_2_, Hb, and lipids in Case 1 obtained by linear unmixing. The scalebar (A) represents 5 mm, all panels except E have equal resolution.

There is marked improvement over the original OA image (Fig. 1C), reconstructed using a uniform speed-of-sound (SoS) model-based inversion,^55^ when compared to the image afforded by applying all the steps of the image formation pipeline proposed herein (Fig. 1D). The stepwise improvement achieved (Fig. 1E) is shown for the region of interest (ROI) delineated in Fig. 1C, D with a dotted rectangle. Each step improves the image, minimizing background noise, the appearance of ring artifacts, and several other distortions, resulting in significantly higher image fidelity.

Even typical model-based inversion makes use of assumptions on sound propagation and detector properties that do not accurately capture the physical parameters of the experimental measurements. The use of a dual speed-of-sound model improves spatial distortion and better reconstructs the position of a vascular structure, while incorporation of the total impulse response,^58^ signal averaging, and spectral median correction reduces ring artifacts and improves the resolution, signal-to-noise, and contrast-to-noise ratios (Fig. 1E). Fig. 1F summarizes the contrast-to-noise (CNR) improvements afforded by each step across all 22 images from the study; an overall mean CNR improvement of 3.6 dB is achieved. Marked resolution improvements were observed after TIR correction (Fig. 1G), resulting in a sharper appearance of blood vessels and other image features. TIR correction can yield an image resolution near those dictated by the detector hardware;^58^ here the CNR was improved by an average of ∼2.6 dB. Frame averaging^59^ (3-frames; see Methods 4.1.3) resulted in a further mean CNR improvement of 0.9 dB, using elastic image registration to reduce blurring due to motion by aligning successive reconstructed images prior to averaging. Ring-shaped artifacts caused by electrical noise (Fig. 1E, H; yellow arrows) were suppressed by spectral median processing, whereby every pixel was replaced by the median of its spectrum in a narrow wavelength range (see Methods 4.1.4). Spectral median correction exploits the premise that the absorption of endogenous chromophores varies smoothly with illumination wavelength, whereas noise appears as peaks at arbitrary wavelengths.

A different representation of the improvements can be seen in the dual band rendering (Fig. 1I). Such images can be shown by combining reconstructions performed on raw data under different high-pass filtering. While the image shown in Fig. 1D was produced using a high-pass filter band with a cut-off frequency of 700 kHz (LCO_700_; see Methods 4.1.1), the use of data containing lower frequencies (Fig. 1J; 100 kHz) allows the reconstruction of lower spatial frequency features. However, the stronger signal at lower frequencies obscures the fine details in the images, as seen in Fig. 1D. Therefore, dual-band representation affords a better representation of both larger structures (in red) and finer structures (in yellow), providing an optimal display of optoacoustic information within the ultrasound bandwidth recorded by 2G-OPUS.

The *Acuity Echo®* scanner used in this study features a fast-tunable laser (25 Hz, i.e., up to 25 different wavelengths/sec.), allowing recording of images in the 700–970 nm wavelength range at 10 nm intervals. Images at different wavelengths exhibit sensitivity to different tissue chromophores and provide thereby molecular information (Fig. 1K), in particular deoxyhemoglobin (Hb), oxyhemoglobin (HbO_2_), and lipids.^60^ To examine the relative appearance of OA human breast images at different wavelengths, we also rendered images at 700 nm (Fig. 1L), 880 nm (Fig. 1M), and 930 nm (Fig. 1N). Using linear unmixing, the contributions of different chromophores can be also resolved and displayed through color-coding (Fig. 1O). This rendering shows the relative bio-distribution of the components captured by the OA method, adding functional and tissue composition information to morphological images obtained by US and single-wavelength OA images. The image shows the subcutaneous vasculature and vascular features around the tumor, color-coding red shades for oxygenated hemoglobin (HbO_2_) and blue shades for deoxygenated hemoglobin (Hb), while adipose tissue is shown in green. Only the upper edge of adipose tissue is shown, due to the applied high-pass signal filtering, which suppresses bulky structures. The melanin layer in the skin also appears in blue, due to the similarity of the melanin absorption spectrum to that of Hb.

### 2.2 Characterization of breast cancer features

Following the demonstration of improved image quality and fidelity achieved by using curved detector arrays and model-based inversion, we aimed to identify the key OA features of breast tumors. We inspected OA images from 22 scans of breast lesions (see Methods, 6), grouped the observations that correspond to malignancy, and linked the findings to histological analysis and the clinical description of the lesions imaged.

To exemplify our findings, Figure 2 showcases three representative cases (Cases 1–3; see Table 1). Images obtained from Case 1 (Fig. 2A–D; used also for the demonstration of image improvement in Fig. 1), identified elongated blood vessels, up to 2 mm in diameter, starting more than 1 cm away from the lesion and centripetally arranged around the tumor mass. Dual-band visualization (Fig. 2A) showed high vascular density surrounding the tumor core. Moreover, vascular contrast in a patchy distribution is seen within the tumor mass. Histological analysis identified a lesion with a well-defined central core and spiculated infiltration typical of invasive lobular carcinoma (Fig. 2C). Vasodilation in the tumor neighborhood was histologically confirmed (Fig. 2D). The appearance of the OA image is markedly different than that of the ultrasound, showing involvement of a larger part of the breast tissue compared to the ultrasound-based characterization of the tumor extent (Fig. 2B; grayscale).

**Figure 2.**
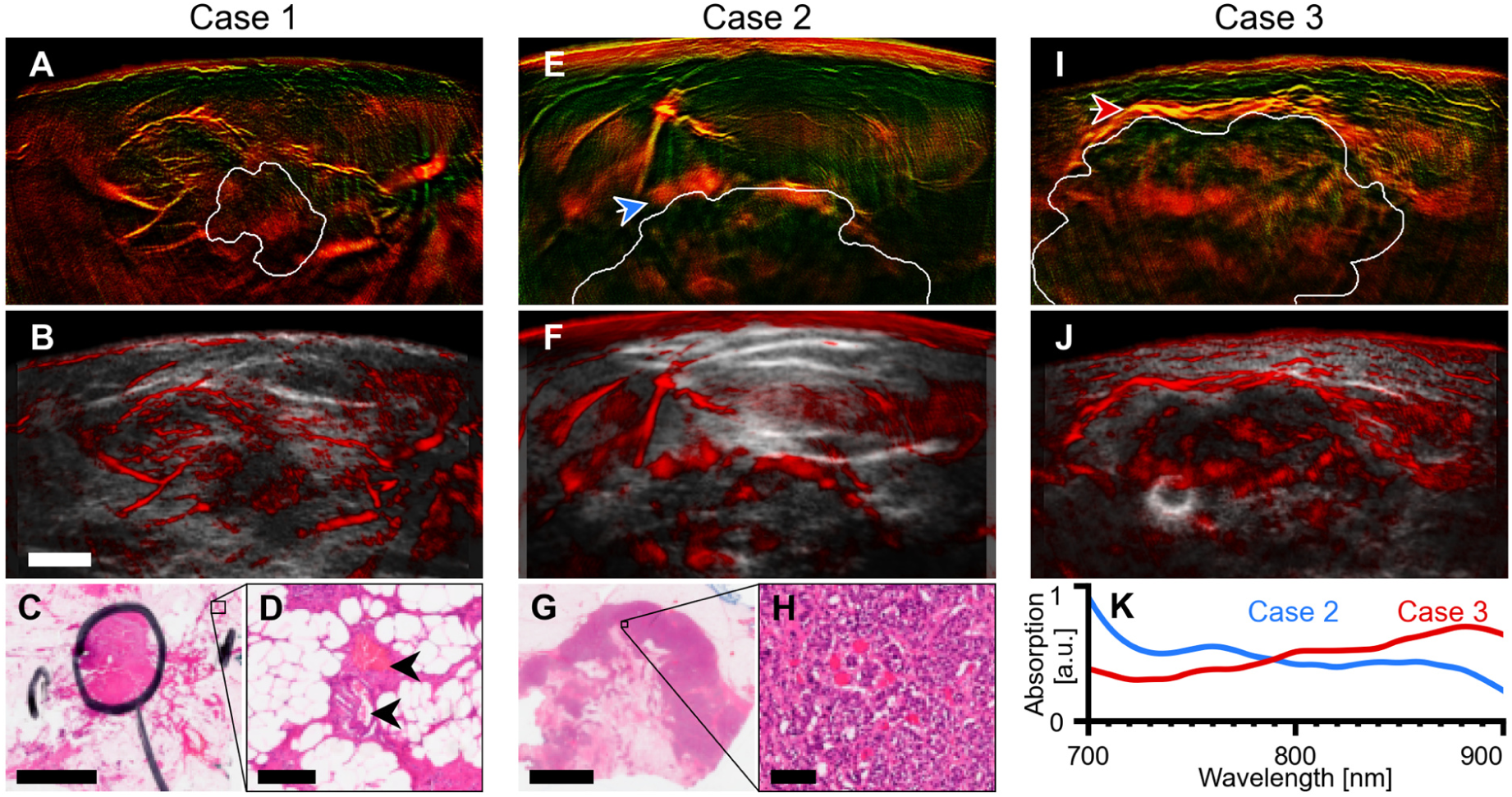
MSOT visualizes vascularization and perfusion in the tumor area at high resolution. **A–D**, Case 1, an invasive lobular carcinoma. Dual-band visualization (A) and OPUS visualization (B) of the median of images in the 850–890 nm range shows rich vasculature around the tumor core (white contour, segmented manually in the ultrasound image). Histopathology of the excised tumor (B; H&E stain) revealed that this pattern is likely caused by vasodilation induced by cancer cell infiltration outside of the main core (black pen circle). Magnification cut-out (C) shows blood vessels (black arrows) outside the tumor core surrounded by cancer cells. Whereas grayscale ultrasound underestimates the true tumor extent, 2G-OPUS provides additional information about the true cancer spread into the surrounding tissue. **E**–**H**, Case 2, a hormone-positive mamma carcinoma NST. Dual-band visualization (E) and OPUS visualization (F) of the median of images in the 700–740 nm range shows a markedly increased signal in the upper rim of the tumor. Histology slice (H&E stain) of the excised tumor (G) shows a necrotic center of the mass (result of an earlier therapy) and a region of densely concentrated tumor cells at the lesion upper rim. The high-resolution cut-out (H) shows that the rim region is highly perfused by a dense network of tiny capillaries. The individual capillaries are too small for 2G-OPUS to resolve; instead, the increased perfusion is exhibited as a patch of stronger signal. **I**,**J**, Case 3, a HER2-positive NST mamma carcinoma. Dual-band visualization (I) and OPUS visualization (J) of the median of images at 850–890 nm reveals marked increase in vascularization in the upper tumor rim. Contrary to the Case 3 shown in (E), the vessels in this tumor rim are larger and can be clearly recognized in the 2G-OPUS image. **K**, Comparison of mean absorption spectra of the enhanced rims in Cases 2 and 3 (regions denoted by arrows in E and I). The spectrum of rim of Case 2 has a notable signature of deoxyhemoglobin (cf. Fig. 1K), which indicates reduced function of the capillary network. This observation correlates with the patient’s lack of response to neoadjuvant chemotherapy (cT2→ypT3). On the other hand, the spectrum of rim of Case 3 exhibits higher oxygen saturation and suggests better vascular function in supplying the tumor microenvironment. This correlates with this patient’s positive response to neoadjuvant chemotherapy (cT2→ypT1c). Scalebars represent 5 mm (B, C, G), 200 μm (D), and 100 μm (H).

**Table 1:** Overview of the cases described in detail herein (Figures 2, 3, and S1). Size denotes the diameter of the hypoechoic tumor core in the selected image, depth refers to the distance between the tumor center of mass and the skin surface. Estrogen receptor (ER) and progesterone receptor (PR) statuses are reported in percent, except Case 4 which was rated by a different histopathologist and is reported as immunoreactive score. All patients were female. IHC = immunohistochemistry.

| Case | Age |  | Size (cm) | Depth (cm) | Grade | ER | IHC |  | HER2 |
| --- | --- | --- | --- | --- | --- | --- | --- | --- | --- |
|  | range | Lesion type |  |  |  |  | PR |  |  |
| Case 1 | 71-75 | ILC | 1.5 | 1.3 | 1 | 100 | 100 |  | Neg. |
| Case 2 | 61-65 | No special type | 3.1 | 2.2 | Gx | 70 | 10 |  | Neg. |
| Case 3 | 51-55 | No special type | 3.2 | 1.6 | 2 | 9/12 | 6/12 |  | Pos. |
| Case 4 | 76-80 | No special type | 0.8 | 0.8 | 3 | 3 | 0 |  | Pos. |
| Case 5 | 26-30 | Fibroadenoma | 1.7 | 0.8 | N/A | N/A | N/A |  | N/A |
| Case 6 | 46-50 | No special type | 1.1 | 2.3 | 2 | 10 | 0 |  | Neg. |
| Case 7 | 46-50 | No special type | 0.8 | 0.8 | 2 | 100 | 100 |  | Neg. |
| Case 8 | 61-65 | Inflammatory | 1.1 | 2.1 | 2 | 10 | 0 |  | Neg. |

Case 2 (Fig. 2E–H) is a hormone-positive mamma carcinoma of no special type (NST). Histology identified a necrotic core and a highly perfused rim region with high cancer cell density (Fig. 2G, H) and confirmed the OA appearance showing high vascular density around the tumor rim (Fig. 2E, F); one such area is highlighted with an arrow on Fig. 2E. Case 3 (Fig. 2I, J) is also shown to better illustrate the carcinoma pattern seen in Case 2 and depicts a HER2-positive NST tumor with a markedly vascularized upper rim (Fig. 2I arrow). MSOT reveals a dense vascular bed surrounding the tumor mass. Fig. 2K compares the absorption spectra of the rims in Cases 2 and 3. Case 2 has an absorption spectrum dominated by deoxyhemoglobin, whereas Case 3 shows better oxygenation. This observation suggests that the vasculature in the rim of Case 3 is more functional than in Case 2. Following the MSOT scans, both patients underwent neoadjuvant chemotherapy. The Case 2 patient did not exhibit any response (increase from cT2 to ypT3), whereby Case 3 had a positive response (decrease from cT2 to ypT1c). This finding can be explained by previous observations showing that tumor oxygenation correlates with neoadjuvant chemotherapy outcome.^61,62^

Figure 3 shows Cases 4–6, which demonstrate additional characteristic patterns revealed on the OA images. Case 4 (Fig. 3A–D) shows a HER2-positive mamma carcinoma NST with irregularly shaped peritumoral structures seen on the OA images (Fig. 3A, B, arrows 1–3). These peritumoral structures were found to contain increased hemoglobin signal (see Fig. 3E) compared to small blood vessels (Fig. 3A, arrow 4) and to background tissue (Fig. 3A, circle 5). To perform this comparison between tumor and background hemoglobin signals, we selected lesions that were approximately the same depth to minimize possible depth-related attenuation effects on the OA signal. Histology results (Fig. 3C) revealed the presence of numerous pre-cancerous ductal proliferations (black arrows) around the invasive tumor (dotted ellipse), accompanied by chronic periductal inflammation causing edema and increased blood perfusion, as also seen at magnification (Fig. 3D). The presence of the inflammation foci explains our observations of the peritumoral structures described above, as their sizes and spatial arrangement match, and increased hemoglobin concentration has been previously observed in chronic inflammation of the gut.^47^

**Figure 3.**
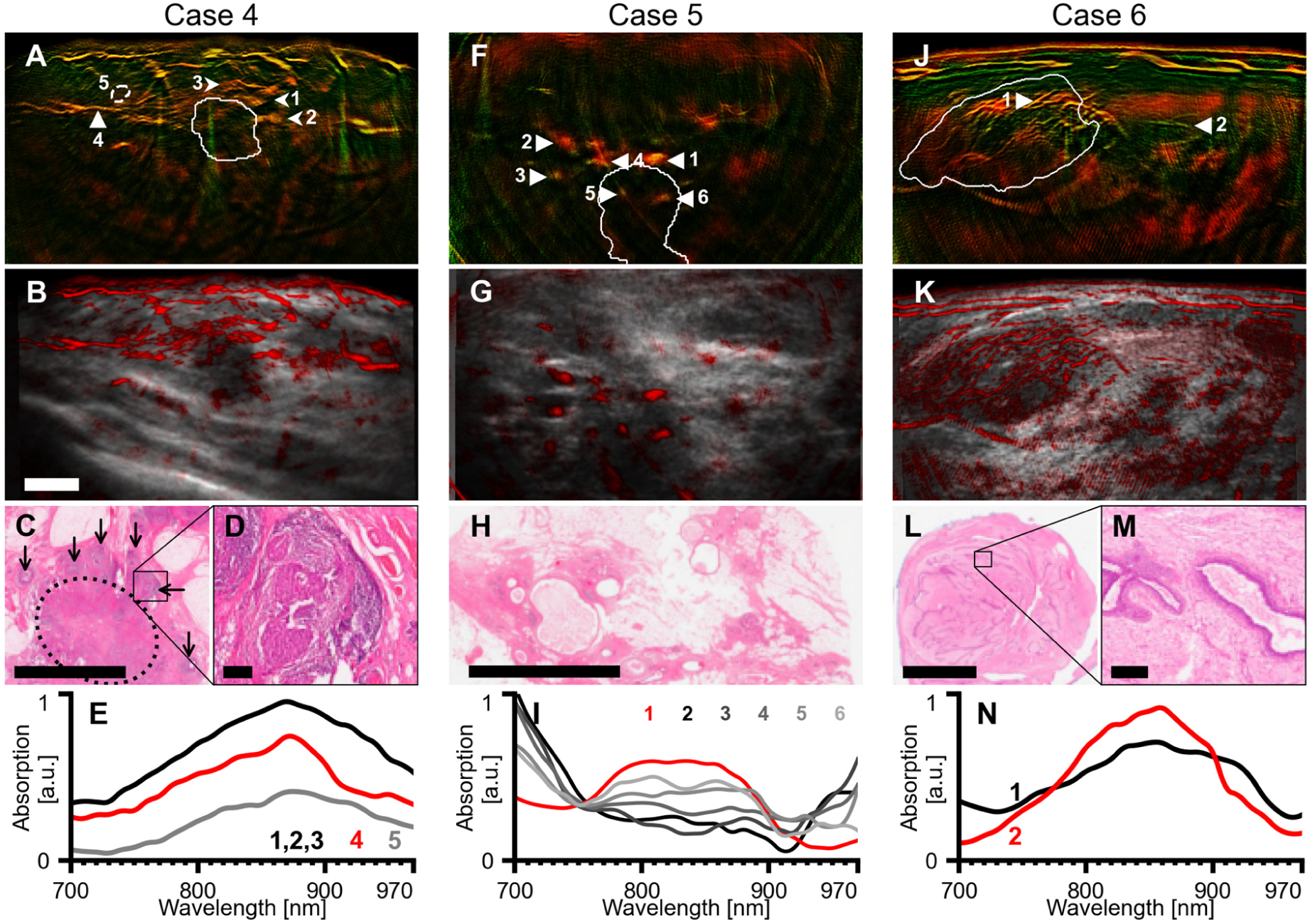
2G-OPUS visualizes signs of inflammation and benign features. **A–D**, Case 4, a HER2-positive mamma carcinoma NST. Dual-band (A) and hybrid OPUS (B) visualizations of median of images in the 850–890 nm range show numerous structures with irregular shape in the vicinity of the tumor core (arrows 1–3). Histology slice (H&E stain) of the excised tumor (C) shows extensive pre-cancerous proliferation in the ducts (thin arrows) around the invasive tumor (dotted ellipse). The high-resolution cut-out (D) shows accompanying chronic periductal inflammation causing edema and increased blood perfusion. **E**, comparison of mean absorption spectra of the inflammation foci (1–3), a small blood vessel (arrow 4), and background tissue (circle 5). **F–H**, Case 5, an ER-positive mamma carcinoma NST. Dual-band (F) and hybrid OPUS (G) visualizations of median of images at 700–730 nm show numerous spots of increased signal around the tumor site. Histology slice (H&E stain) of the excision at the tumor site (H) shows numerous cystically dilated ducts filled with proteinaceous fluid and foam cells. **I**, comparison of absorption spectra of the six spots marked by arrows in F. Whereas the spectrum of spot 1 resembles hemoglobin and suggests this is either a cross-sectionally imaged blood vessel or a cyst with internal bleeding, spot 2–6 have a markedly different absorption spectrum. **J–M**, Case 6, a fibroadenoma. Dual-band (J) and hybrid OPUS (K) visualizations of median of images in the 700–740 nm range show numerous tubular branching structures inside the tumor core (white contour). Histology slice (H&E stain) of the excised tumor (L) shows a typical pattern for fibroadenomas: dense stromal proliferation with a network of compressed ducts. These ducts match the observed pattern in the optoacoustic images in both size and density. The high-resolution cut-out (M) shows a detail of a compressed duct filled with proteinaceous fluid. **N**, comparison of absorption spectra of the ducts (1) and a blood vessel in similar depth (2; marked by white arrows in J). The duct spectrum is similar to the blood vessel spectrum, which indicates a possible bleeding occurring in relation to a past compression of the mass. No signs of bleeding are present in the histological sample which was excised 7 months after the scanning was performed. Tumor cores in A, F, and J, denoted by white contours, were segmented in co-registered ultrasound images. Scalebars represent 5 mm (B, C, H, L) and 200 μm (D, M). Panels A, B, F, G, J, K have the same resolution.

Case 5 (Fig. 3F–I) shows an ER-positive mamma carcinoma with notable cystic dilation of the ducts around the tumor site visible in the histology slice (Fig. 3H). OA images of this tumor (Fig. 3F, G) show numerous patches of increased signal (arrows 1–6), as opposed to previous observations from tumors. Some of the patches are colocalized with anechoic capsules on the US (arrows 2 and 3). The mean absorption spectra of patches 2–6 (Fig. 3I) are also different than those taken from peritumoral lesions and attain a spectral profile representative of a mixture of deoxyhemoglobin and water, similar to absorption profiles of breast cysts previously reported using optical imaging.^63-65^ This spectral appearance is different from that of blood vessels, which are dominated by oxyhemoglobin signal, as in patch 1. Compared to the optical imaging studies, the high resolution and spectral contrast afforded by our OA image formation pipeline enables more accurate signal interpretation and the differentiation of cysts from blood vessels.

We also showcase MSOT features revealed from a benign lesion—a fibroadenoma (Case 6; Fig. 3J–N). The OA images show numerous tubular structures within the tumor core (Figs. 3J, K). However, it is apparent in the high-resolution image in Fig.3J that the dense vascular network around the rim seen around malignant tumors (Fig. 2) is not present. Histology slices of this tumor show that the observed structures correspond to numerous compressed ducts filled with proteinaceous fluid that form a linear branching network, a pattern often seen in fibroadenomas (Fig. 3L, M). Fig. 3N shows a comparison of the mean absorption spectrum in one of the ducts (arrow 1) and a small blood vessel at similar depth (arrow 2). The similar appearance of both spectra indicates possible compression-induced intraductal bleeding.

Table 2 summarizes the frequency of the observed patterns in the OA images across all the 22 analyzed lesions. Due to the exploratory nature of our analysis, we do not draw direct conclusions regarding the diagnostic value of these patterns.

**Table 2:** Summary of presence of observed optoacoustic patterns in analyzed lesions grouped by the lesion malignancy status. Our study focuses on better understanding of the patterns visible thanks to high spatial resolution and spectral contrast of 2G-OPUS. We do not draw direct conclusions from this study regarding the diagnostic value of these patterns.

| Lesion type | Malignant<br>(N=16) | Benign<br>(N=6) |
| --- | --- | --- |
| Rim enhancement | 5 | 0 |
| Blood vessels > 1 mm | 5 | 2 |
| Blood vessels < 1 mm | 13 | 5 |
| Cysts, ducts | 2 | 3 |

## 3 Discussion

In this study, we developed a new image processing platform to set a new mark in the image quality of handheld optoacoustic breast cancer imaging. 2G-OPUS achieved unprecedented image quality, which—together with spectral contrast and the high resolution—allowed the characterization of OA features in malignant and benign breast tumors. We examined 22 breast lesions and linked observed patterns to available clinical data and micrographs of the excised tissues. We could identify patterns of malignancy, such as enhanced rim and vascular density and functional parameters, in particular oxygenation/hypoxia, and showcased representative examples seen in eight cases.

We demonstrated that using improved image processing tools—reconstruction models with TIR correction, averaging of motion-corrected frames, and color-coded visualization of two frequency bands—we could achieve optoacoustic imaging quality never before demonstrated with a handheld scanner (*Acuity Echo®*). Our approach showed the ability to resolve blood vessels with diameters as small as 200 μm at depths up to 2 cm (see Case 1— Fig. 2 and Case 7—Fig. S1). Comparable imaging quality has been so far reported only with dedicated bed-based scanners *SBH-PACT*^35^ and *PAM-03*,^36^ which nevertheless yielded lower resolution (255 μm and 370 μm, resp.). Unlike stationary imaging systems, handheld optoacoustic imaging can be seamlessly integrated in routine breast ultrasound,^39-41^ improving the information obtained in the imaging session.

Malignant tumors generally exhibited patterns of small blood vessels arranged centripetally around the tumor core, a finding that has been previously described as one of the most reliable OA imaging features to indicate malignancy.^8,9^ Compared to previous studies however, the superior image quality achieved herein facilitated detailed visualization of this feature, which could increase the confidence in diagnosis of suspicious borderline lesions, thus reducing the number of unnecessary biopsies. In a smaller number of cases, the findings also demonstrated the presence of dilated vasculature surrounding the tumor core. In Case 1, examination of the tumor histopathology revealed that the vasodilation was caused by infiltration of the cancer cells into the surrounding tissue. Whereas standalone US often underestimates the size of ILC,^66,67^ our results show, in line with earlier *ex vivo* experiments,^68^ that optoacoustics can help with assessment of tumor margins *in vivo*. 2G-OPUS could also characterize functional parameters associated with the tumor rim microenvironment. In Cases 2 and 3 we observed different oxygenation levels in the rim, reflecting the functional condition of these vessels, and corresponding differences in neoadjuvant chemotherapy (NAT) response. Compromised functionality, seen as hypoxia, reduces the efficiency of therapeutic drug delivery^69,70^ and has been shown to correlate with poor NAT outcomes.^61,62^ This observation indicates a potential for 2G-OPUS to offer detailed functional images serving as a predictor for NAT outcomes.

We further observed features associated with benign lesions, such as cysts, and chronic inflammation around ducts containing pre-cancerous proliferations. Although the relationship between inflammation and invasive progression of ductal carcinoma in situ (DCIS) is not fully understood,^71^ some studies show that the presence of chronic inflammation relates to increased risk of recurrent invasive disease.^72,73^ MSOT has been used to monitor chronic inflammation of the gut;^47,74^ however, this is the first time it has been used to visualize chronic inflammation in the human breast. Although the inflammation could not be distinguished from malignancy based on functional features such as oxygen saturation, a combination of 2G-OPUS with traditional modalities could facilitate identification of such risky DCIS cases early and could support better treatment decisions.

We found that motion may have a negative effect on the quality of the images and may distort spectral information. The negative influence of motion could be suppressed by shorter scanning time, possibly through recording less wavelengths. Since the current wavelength selection is very dense compared to smooth changes of absorption spectra of the main chromophores, it produces partially redundant information and could be reduced without compromising the spectral information. A future goal is therefore to accelerate the computation speed of the developed processing pipeline to make it available to the operator in real time.

Overall, the combination of improved image processing with handheld, hybrid optoacoustic-ultrasound acquisition, label-free contrast, and fast operation make 2G-OPUS well suited for incorporation into the established breast cancer examination workflow. It expands the capabilities of routinely used conventional grayscale ultrasound with the ability to image additional pathophysiological features. Simultaneous hybrid acquisition of ultrasound along with optoacoustic images allows easy localization of tumors during scanning and eliminates the need for two separate examinations, thus increasing patient comfort. Additionally, the rich spectral information provided by 2G-OPUS imaging allows label-free resolution of endogenous chromophores absorbing in the near-infrared range, primarily oxy- and deoxyhemoglobin, which can be used to estimate oxygen saturation of blood. Since 2G-OPUS can visualize these features within the established clinical routine by expanding the capabilities of common grayscale ultrasound, we see great potential for its translation into standard clinical breast care.

## 4 Materials and Methods

### 4.1 Optoacoustic image processing pipeline

Fig. 4 summarizes the data processing pipeline utilized in this work.

**Figure 4.**
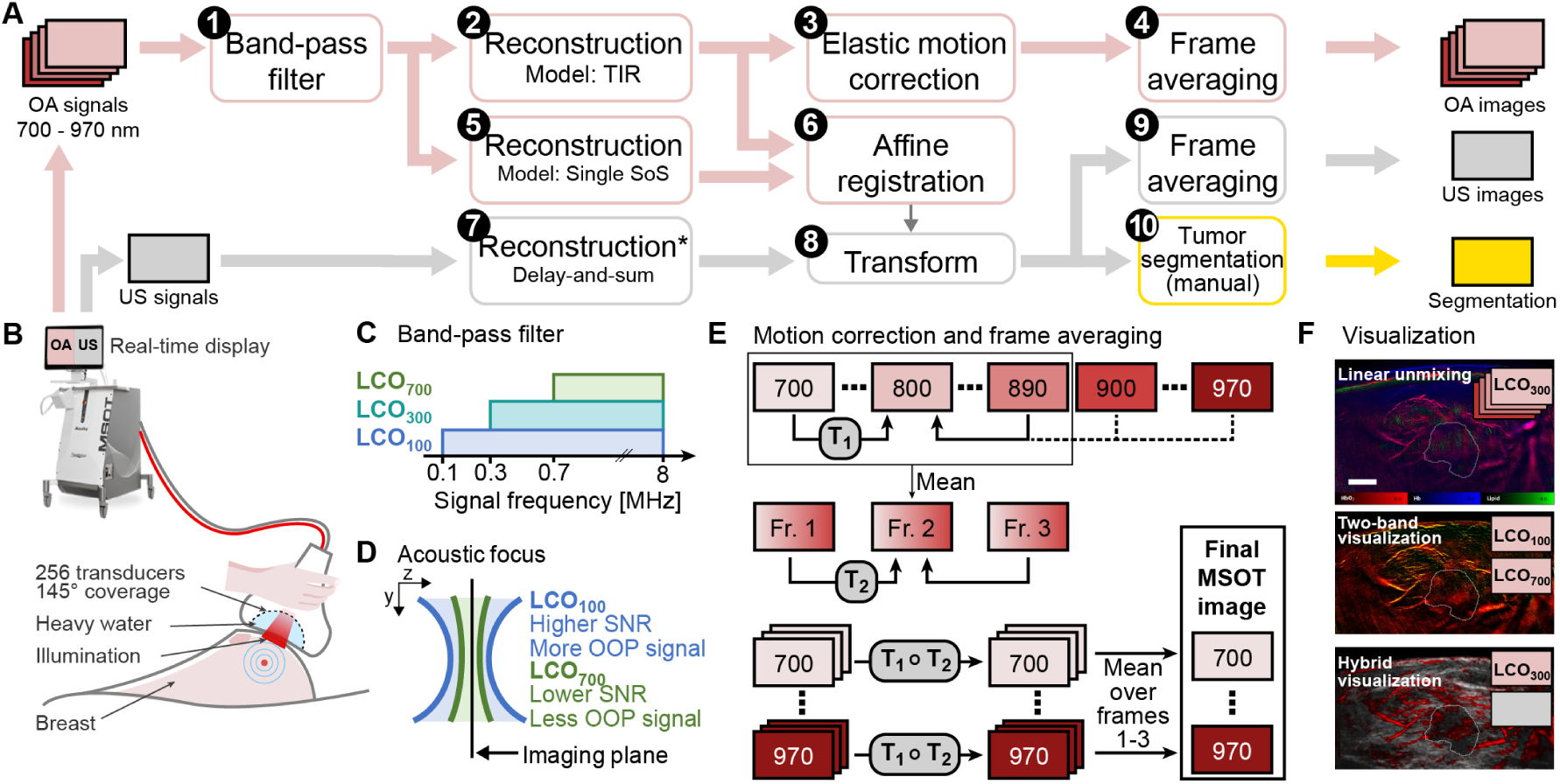
Overview of the data acquisition, processing, and visualization methods used. **A**, Summary of the steps used to process the data off-line: 1, Band-pass is applied to the optoacoustic (OA) signals. 2, Images are reconstructed from the filtered signals using an improved model with total impulse response correction and dual speed-of-sound distribution. 3, Intra- and inter-frame elastic motion correction is applied. 4, Final OA images are produced by averaging the motion corrected frames. 5, OA signals are also reconstructed using a single speed-of-sound model. 6, The two variants of OA images are affinely registered to estimate the distortion caused by variable speed-of-sound models. 7, Ultrasound (US) images are reconstructed by the scanner, assuming a uniform speed-of-sound, as in step 5. 8, US images are transformed using the transformation estimated in step 6 to match the spatial distribution of dual speed-of-sound OA images. 9, Consecutive US images matching the timeframe of the OA images are averaged. 10, Tumors are manually segmented in the US images. **B**, Visualization of the scanner and the handheld probe during the data acquisition. US transducers are arranged in a 145° arc. The arc cavity is filled with heavy water. Laser light is delivered through an optical fiber bundle and a diffuser. US and OA images are displayed on the scanner screen in real time. **C**, Three band-pass filter configurations are used in the step A.1, differing in their lower cut-off: 100 kHz (LCO_100_, 0.1–8 MHz), 300 kHz (LCO_300_, 0.3–8 MHz), and 700 kHz (LCO_700_, 0.7–8 MHz). Each is used for different visualization. **D**, The acoustic focus of the transducers varies with the signal frequency, the out-of-plane signals are more present in the low frequency component of the signal. The band-pass filter (A.1) thus presents a trade-off between suppressing the out-of-plane signals and decreasing the signal-to-noise ratio. **E**, Overview of the motion correction and the frame averaging applied in the steps A.3 and A.4. First, images at wavelengths 700–890 nm are registered to the image at 800 nm (intra-frame elastic displacements T_1_) and their mean is taken to represent the multispectral frame. Three consecutive multispectral frames are then registered to the middle frame (inter-frame elastic displacements T_2_). Images at wavelengths above 890 nm cannot be reliably registered to 800 nm because they are visually very different; T_1_ of 890 nm is used for them instead. All images are then transformed by composite transformations T_1_∘T_2_, and a mean multispectral frame is computed by averaging corresponding single-wavelength images from the three frames. **F**, The resulting multispectral images can be visualized as chromophores using standard linear unmixing. The dual-band visualization can provide an optimal trade-off between out-of-plane signal suppression and low noise in single-wavelength images. Alternatively, a hybrid visualization combining the US and OA images provides tumor features from both modalities together. All visualizations use tumor core contours manually segmented in the co-registered US images.

#### 4.1.1 Band-pass filtering

First, we processed the recorded acoustic signals using the Butterworth band-pass filter. The spatial sensitivity of the ultrasound transducers varies with the sound frequency and the receptive field grows as the frequency decreases, as shown in Fig. 4D. Excluding the low frequencies leads to better acoustic focus on the imaging plane by suppressing out-of-plane signals. However, the OA signals are broadband by their nature and filtering makes the signal incompatible with the physical model. Removing low frequencies thus introduces artifacts and results in noisier images. To explore these effects in detail, we used three different variants of the Butterworth filter (8^th^ order low-pass and 2^nd^ order high-pass) and selected the appropriate one for each visualization. The higher cut-off value was set to 8 MHz to match the transducer sensitivity; for the lower cut-off (LCO) value we used three increasing thresholds (Fig. 4C): 100 kHz (LCO_100_), 300 kHz (LCO_300_), and 700 kHz (LCO_700_).

#### 4.1.2 Model-based image reconstruction

Second, we reconstructed the images by computing the initial pressure *p*_0_ from the filtered signals *s* using an iterative model-based approach. We used acoustic model *M* of the scanner which accounts for the different speed of sound in the tissue and in the probe cavity filling (heavy water), the wave refraction on their interface, and for the physical properties of the transducers summarized in the total impulse response (TIR) of the system.^58^ Additionally, we used Tikhonov regularization to address the ill-posedness of the inverse problem and mitigate the limited view artifacts and measurement noise. The regularization parameter *λ* was chosen using an L-curve. A non-negative LSQR algorithm was used to solve the optimization problem:

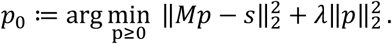

The reconstructed OA images are of the size 401 × 401 pixels and correspond to FOVs of 4 cm × 4 cm.

#### 4.1.3 Elastic motion correction and frame averaging

Finally, sequences of three consecutive multispectral images were co-registered and averaged to reduce the noise (Fig. 4E), using the following two-step procedure. We denote *I*^*n*^[*λ*] a single-wavelength image in a multispectral frame *n* at wavelength *λ* (nm). First, intra-frame elastic transformations 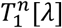 are estimated, aligning images *I*^*n*^[*λ*] to *I*^*n*^[800] for *λ* ∈ [700, …, 890]. The transformation *T*_*1*_ cannot be estimated for images at wavelengths over 900 nm, as these have a very different appearance from the rest of the stack; instead, we set 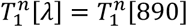 for *λ* ∈ [900,.., 970]. For the second step, we generate mean images *Ī* ^*n*^ to represent individual multispectral frames, as 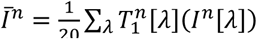, for *λ ∈* [700, …, 890]. Inter-frame elastic transformations 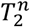 are estimated by aligning mean images *Ī*_*n*_ to *Ī*_2_. Final images *I*\*[*λ*] are created by computing the mean of the co-registered images as 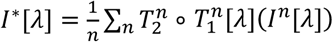, for *n* ∈ [1,2,3]. The registration was performed using the SimpleITK library.^75^

#### 4.1.4 Spectral median adjustment

When considering single-wavelength images, such as in dual-band or hybrid visualizations, we replaced the intensity values in each pixel by median of the pixel spectrum in a narrow wavelength range. We used range of 40 nm, corresponding to 5 wavelengths in our setup: *Ĩ*[*λ*] = med(*I*[*λ* − 20], *I*[*λ* − 10], *I*[*λ* + 10], *I*[*λ* + 20]). Thanks to relatively smooth variation of the absorption spectra of the main endogenous chromophores, this step further helped to improve the image contrast.

### 4.2 Ultrasound image processing

The ultrasound images were reconstructed by the scanner using an algorithm that assumes a homogenous speed-of-sound (Fig. 4A-7), which means they were spatially distorted compared to the OA images. To estimate this distortion, we additionally reconstructed the OA images using a model assuming the same homogenous speed-of-sound model (Fig. 4A-5) and computed an affine transformation between the two variants of OA images (Fig. 4A-6). This transformation was then used to warp the ultrasound images to approximately match their spatial frame to the dual SoS OA images (Fig. 4A-8). Finally, we picked the US frames which were recorded concurrently with the final averaged MSOT frames and averaged them to increase their SNR (Fig. 4A-9).

The tumor cores were manually segmented in the processed ultrasound images in consensus with the performing radiologist (Fig. 4A-10).

### 4.3 Contrast-to-noise ratio computation

We compute contrast-to-noise ratio as

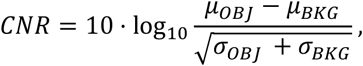

where *μ* and *σ* represent mean and variance, respectively. We define the object as the tissue up to 1.5 cm under the skin and background as the area above the skin. When computing the statistics over our whole dataset, we evaluate the CNR on LCO_700_ variant of the images at 880 nm (or 850–890 nm when considering the spectral median).

### 4.4 Visualization methods

Optoacoustic signal decays with depth considerably due to light absorption and scattering, resulting in images with very high range of intensity values. To improve the contrast in our visualizations, we apply local contrast normalization and sigmoid normalization. Denoting the image as *I:* Ω ⊂ ℝ^2^ ↦ ℝ, the *n-*th percentile of intensities within the image as *P*_*n%*_, and defining a neighborhood of a pixel *x* as *N*_*x*_ = {*x*′ ∈ Ω | ‖ *x* − *x*′‖_1_ ≤ *r*}, local contrast normalization transforms the image as

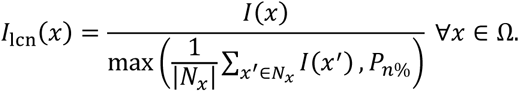

Sigmoid normalization maps the intensities of an image to the interval [0,1], mapping values in the percentile range [*P*_*a*_, *P*_*b*_] to [*a, b*] and squeezing the outside values to the ends of the interval:

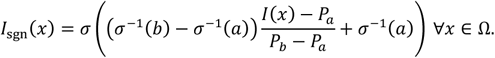

Here, *σ* denotes the logistic sigmoid. Additionally, we apply power law transformation (*I*_pl_(*x*) = *I*(*x*) ^*γ*^) and unsharp masking (*I*_um_ *= I + α (I* − *G*_*σ*_ *\* I*), where *G* _*σ*_ is Gaussian kernel with scale *σ*). We choose the values of parameters *r, n, a, b, γ, α, σ* manually for each image. In case of multi-channel images, these transformations are applied to each channel individually.

Ultrasound images, normalized to the 0–1 range, employ the following depth gain correction and contrast enhancement procedure:

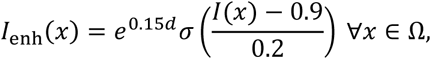

where *d* is the depth of pixel *x* under the tissue surface in centimeters.

Linear unmixing visualization (Fig. 2F) shows coefficients obtained by solving the constrained linear system *Sc* = *p,c* ≥ 0, where *S* are stacked absorption spectra of oxy- and deoxyhemoglobin, fat, and water,^60^ and *p* is the observed spectrum. Coefficient values are processed with the above transformations and displayed as red (oxyhemoglobin), blue (deoxyhemoglobin), and green (fat) color channels. Water coefficient is not displayed.

Dual-band and hybrid OPUS visualizations use 2D colormaps created by interpolation among pre-defined corner values in the RGB color space.

### 4.5 OPUS setup

In-vivo measurements were acquired with an *Acuity Echo®* (iThera Medical GmbH, Munich, Germany) hybrid handheld multispectral optoacoustic and ultrasound scanner (Fig. 4B). Tissue illumination was performed in short light pulses (duration ∼8 ns) that were produced by a tunable laser and delivered to the probe via an optical fiber bundle. A diffuser was used to produce a rectangular illumination spot on the skin (ca. 0.5×3 cm). The peak pulse energy was ≈16 mJ, which is within the permissible energy exposure limits set by the American National Standards Institute.^59^ Ultrasound signals, produced upon absorption of light energy in the tissue through the optoacoustic effect, were received by 256 piezoelectric elements (4 MHz central frequency) arranged to a curved linear array with 145° coverage and 6 cm distance between the endpoints. The cavity inside the arc was filled with heavy water (D_2_O) to ensure acoustic coupling while minimizing absorption of near-infra-red light. We recorded images at 28 separate wavelengths between 700 and 970 nm at 10 nm intervals using a single pulse-per-image acquisition with framerate of 25 Hz; acquisition of a full multispectral OA frame consisting of 28 single-wavelength images took 1.1 s. Pulse-echo ultrasound images were acquired synchronously with the OA images during the pauses between individual laser pulses. The synthetic transmit aperture method with spatial compounding of sub-apertures was used.^76^ The US images were acquired at the repetition rate of 6.25 Hz.

Both the US and the OA images were displayed in real-time on the screen of the scanner. These images were reconstructed using delay-and-sum algorithm, which can run in real-time but produces images of inferior quality compared to off-line model-based reconstructions. To enable high-quality off-line reconstructions, the raw optoacoustic signals were stored.

### 4.6 Dataset

The aim of this clinical study was characterization of the features extracted by hybrid 2G-OPUS imaging from breast tumor tissue using the 2G data processing pipeline. We employed scans from 22 female patients (age range 20’s–70’s) with breast tumors that were clearly visible in the US and the OA images and were accompanied with complete information for each scan. The scans were performed by a senior radiologist experienced in breast ultrasonography who received additional training for 2G-OPUS imaging. The study was approved by the local ethics committee of the Technical University of Munich (Nr. 27/18 S) and all participants gave written informed consent upon recruitment. The tumor types scanned are summarized in Table 3.

**Table 3:**
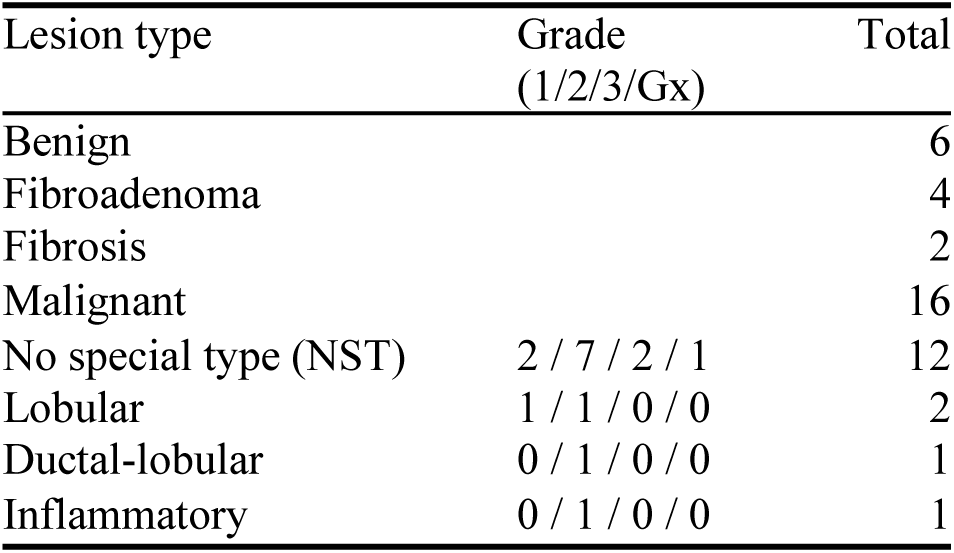
Overview of the analyzed masses by their type and cancer grade distribution.

| Lesion type | Grade<br>(1/2/3/Gx) | Total |
| --- | --- | --- |
| Benign |  | 6 |
| Fibroadenoma |  | 4 |
| Fibrosis |  | 2 |
| Malignant |  | 16 |
| No special type (NST) | 2 / 7 / 2 / 1 | 12 |
| Lobular | 1 / 1 / 0 / 0 | 2 |
| Ductal-lobular | 0 / 1 / 0 / 0 | 1 |
| Inflammatory | 0 / 1 / 0 / 0 | 1 |

Patients were scanned in the supine position in a quiet room with normal temperature (≈23°C). Standard ultrasound gel was used to ensure acoustic coupling between the probe and the tissue. Anatomical navigation and selection of the desired field-of-view was achieved thanks to simultaneous co-registered ultrasound imaging provided by the hybrid 2G-OPUS setup. Each tumor was scanned for several seconds to record multiple frames. Nevertheless, the patients were not required to hold their breath during the examination period. The total examination length did not exceed 15 minutes, including patient preparation.

Irrespective of the study participation, all patients underwent surgical removal of the tumor as part of the planned treatment. Histology images shown herein were obtained from the excised, paraffin-embedded tissue and stained with hematoxylin-eosin for standard histopathological analysis. Evaluation of the images for our study was performed retrospectively by a senior pathologist.

## Data Availability

The data that support the findings of this study are available on request from the corresponding author. The data are not publicly available due to privacy or ethical restrictions.

## Acknowledgements

This project has received funding from the European Research Council (ERC) under the European Union’s Horizon 2020 research and innovation programme under grant agreement No 694968 (PREMSOT), by the Deutsche Forschungsgemeinschaft (DFG), Sonderforschungsbereich-824 (SFB-824), subproject A1 and by the Helmholtz Association of German Research Center, through the Initiative and Networking Fund, i3 (ExNet-0022-Phase2-3).

## Ethics Approval and Consent to Participate

The study was approved by the local ethics committee of the Technical University of Munich (Nr. 27/18 S) and all participants gave written informed consent upon recruitment.

## Abbreviations

CNR: Contrast-to-noise ratio
DCIS: Ductal carcinoma in situ
FWHM: Full width at half maximum
ILC: Invasive lobular carcinoma
LCO: Lower cut-off
MSOT: Multispectral Optoacoustic Tomography
2G-OPUS: 2nd generation Multispectral Optoacoustic-Ultrasound Tomography
NAT: Neoadjuvant chemotherapy
NST: No special type
OA: Optoacoustics
TIR: Total impulse response
US: Ultrasound
SoS: Speed-of-sound

## 5 Supplementary material

### Additional cases

Case 7 (Fig. S1A–D) is a hormone-positive mamma carcinoma of no special type. MSOT image of the tumor (Fig. S1A) as well as hybrid OPUS image (Fig. S1B) exhibit small vessels in centripetal arrangement around the tumor core. Histopathology (Fig. S1C, D) confirms the presence of numerous vessels (arrows) around the tumor (ellipse) with sizes matching our optoacoustic observations. Case 8 (Fig. S1E, F) is an instance of inflammatory breast cancer (IBC; Case 8). Although IBC is not an actual inflammation, it is typically accompanied by symptoms resembling acute mastitis, including skin thickening and redness. MSOT captures both symptoms: the cutis layer is considerably denser than in other scans (cf. Figs. S1A, 2E, 3J) and conspicuous dilation of subcutaneous vasculature correlates with observed erythema.

**Figure S1.**
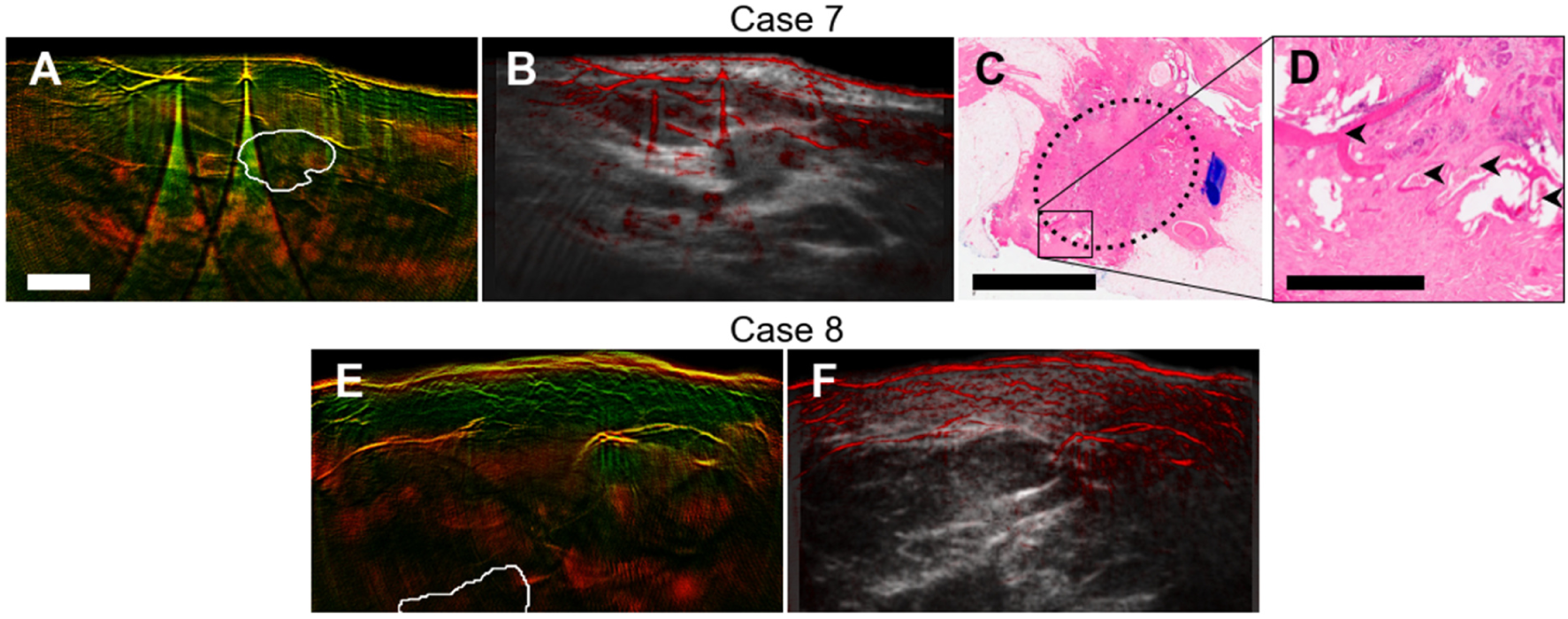
Additional cases of optoacoustic features of breast cancer. **A–D**, Case 7, a HER2-negative mamma carcinoma NST. Dual-band visualization (A) of median of images in the 850–890 nm shows centripetal blood vessel arrangement around the hypoechoic core (segmented in co-registered ultrasound image (B), white contour). Histopathology of the excised tumor (C; H&E stain; marked by dotted ellipse) confirmed presence of numerous vessels (arrows in D) adjacent to the tumor with sizes corresponding to OA observations). **E, F**, Case 8, an inflammatory mamma carcinoma. Dual-band visualization of median of images at 850–890 nm shows thickened cutis and conspicuous dilation of subcutaneous vasculature. This is related to the inflamed appearance of the skin accompanying the tumor proliferation. White contour delineates the tumor core, segmented in a co-registered ultrasound image (F). The scalebar represents 1 mm in F and 5 mm in A and C. Panels A, B, E, F have the same resolution.

